# Gut microbiome associations with incident cardiovascular disease in Nordic populations

**DOI:** 10.64898/2026.11.12.26367973

**Authors:** Brooke N. Wolford, Susanna C. Larsson, Maria Nethander, Lina Lawenius, Kamalita Pertiwi, Oleg Kambur, Eivind Coward, Mari N. Gynnild, Guro F. Giskeødegård, Bjørn Olav Åsvold, Louise Grahnemo, Burak Adnan Erkorkmaz, Gunnar Engström, J. Gustav Smith, Veikko Salomaa, Koen Dekkers, Johan Ärnlöv, Aki Havulinna, Camila Gazolla Volpiano, Guillaume Méric, Teemu Niiranen, Marju Orho-Melander, Tove Fall, Claes Ohlsson, Kristian Hveem

## Abstract

**Background:** The gut microbiome is an emerging risk factor for cardiovascular disease. However, there is a lack of large-scale metagenomic studies with replication that identify gut microbial features associated with incident cardiovascular disease (CVD).

**Methods:** We tested for gut microbiome associations with CVD using metagenomes from stool samples in a discovery-replication framework across four large prospective Nordic cohorts with 635 incident CVD in 11,604 participants in the discovery cohort and 2,309 incident CVD in 20,965 participants combined across three replication cohorts.

**Results:** Higher Shannon diversity index and species richness were consistently associated with a lower risk of incident CVD. In addition, the relative abundance of nine species and eleven bacterial functional modules showed reproducible associations with incident CVD in the discovery cohort and in a meta-analysis of the three replication cohorts. We identified a female-specific gut microbiota-related pathway involving the prevalent oral–gut species *Rothia mucilaginosa_A*, enriched among denture users, thus serving as a proxy for longstanding poor oral health. Mendelian randomization analysis supported a causal effect of denture use on increased relative abundance of *Rothia mucilaginosa_A*. This pathway was linked to increased CVD risk through altered gut microbial manganese transport, elevated circulating imidazole propionate levels, and increased levels of systemic inflammation biomarkers. The second pathway involved both α-diversity metrics and the remaining eight species, all of which were strongly associated with multiple markers of metabolic syndrome and with bacterial pathways related to purine metabolism, microbial stress responses, altered redox balance, ascorbic acid degradation, and branched-chain amino acid biosynthesis.

**Conclusions:** These findings demonstrate that gut microbiota diversity, specific taxa, and functional pathways provide new insights into the underlying biology of CVD. Furthermore, these findings suggest that the link between poor oral health and cardiovascular disease may, at least partly, operate through altered gut microbiota.

## Introduction

Cardiovascular disease (CVD) is the leading cause of morbidity and mortality worldwide^1^. Despite substantial advances in prevention and treatment, considerable heterogeneity in individual risk persists, underscoring the need to identify novel biological pathways that contribute to disease development.

Increasing evidence suggests that the gut microbiome, a complex community of microorganisms, may contribute to cardiovascular health and disease^2^. In a cross-sectional analysis in the Framingham Heart study, decreased overall gut microbial α-diversity, determined by 16S rRNA gene sequencing, was associated with an increased calculated 10-year risk of CVD^3^. In a Chinese cohort, a gut bacterial co-abundance group was associated with coronary artery disease severity^4^ and in a Dutch cohort, four gut microbial species were associated with cardiovascular risk^5^. Finally, in a cross-sectional analysis, higher gut relative abundances of *Streptococcus* species were associated with subclinical coronary atherosclerosis in the Swedish SCAPIS cohort, which was included as one of the replication cohorts in the present study^6^.

These relationships between the microbiome of the gastrointestinal tract and cardiovascular health are referred to collectively as *the gut–heart axis* and are thought to be mediated, at least in part, by gut microbiota–derived metabolites^7^. One such metabolite is trimethylamine N-oxide (TMAO), for which elevated circulating plasma levels have been associated with an increased risk of incident CVD^8,9^. Another metabolite, imidazole propionate (ImP), a histidine-derived molecule produced by the gut microbiota, has been linked to atherosclerosis^10,11^. Furthermore, CVD has been linked to decreased abundance of gut microbial species that produce butyrate^12–18^. Butyrate and other short-chain fatty acids, which are end products of dietary fiber fermentation by specific gut bacterial species, have been shown to exert beneficial effects on pathways implicated in the regulation of cardiovascular disease^19^.

Despite these recent studies, the broader landscape of gut microbial features associated with incident CVD events remains incompletely characterized, and there is a lack of large-scale metagenome sequence-based investigations with successful replication of associations between gut microbial features and incident CVD. In this study, we investigate the associations between gut microbiota α-diversity metrics, relative abundance of metagenomic species, and metagenomic functional pathways and incident CVD across four large, well-characterized, longitudinal population-based Nordic cohorts with stool-derived metagenomic sequencing. We aimed to identify microbiome signatures that may inform biological mechanisms and identify potential pathways for intervention. To our knowledge, this is the first metagenomic study to identify consistent associations between gut microbiome features and incident CVD within a discovery–replication framework.

## Methods

### Discovery cohort — the Trøndelag Health Study

We used data from the fourth survey of the population-based Trøndelag Health Study (the HUNT Study) in Norway as the discovery cohort. The HUNT Study was first conducted in 1984-86 (HUNT1) with surveys taking place roughly every ten years until HUNT4 (2017-2019)^20–22^. Individuals over age 20 living in Trøndelag county were invited to take part in different assessments consisting of questionnaires, interviews, clinical examinations, laboratory measurements and providing biological samples^23^. International Classification of Diseases (ICD) diagnosis codes were obtained from inpatient and outpatient specialist care records at St. Olav’s University, Namsos, and Levanger Hospitals, which together provide specialist and emergency care for the population covered by the HUNT study and biobank. Self-reported dietary intake, alcohol consumption, and smoking were defined as described in the Supplementary Methods. The use of proton pump inhibitors (ATC code A02BC) or antibiotics (ATC code J01 and J04 if available) in the 3 months prior to sampling was collected from the national pharmaceutical registry and used as binary variables.

### Replication cohorts

We externally validated our findings in three Nordic population-based cohorts (Figure 1, Table 1). The Swedish CArdioPulmonary bioImage Study (SCAPIS) is a prospective study that invited a random selection from the population register of individuals aged 50-64 living in 6 counties in Sweden^24^. The metagenomics data from SCAPIS comes from the Uppsala and Malmö sites^25^. The Swedish Infrastructure for Medical Population-based Life-course and Environmental Research (SIMPLER) comprises two longitudinal cohorts (the Swedish Mammography Cohort and the Cohort of Swedish Men), from which a subset with available metagenomic data was included in the present analyses^26,27^. FINRISK is a population-based health survey and the 2002 cohort is a stratified random sample of adults aged 25-74 from 6 regions in Finland with metagenomics^28–30^. Detailed descriptions of these cohorts are available in the Supplementary Methods. As dietary covariates differed between cohorts, each cohort handled these covariates in as harmonized manner as was possible, as is described in the Supplementary Methods.

**Figure 1.**
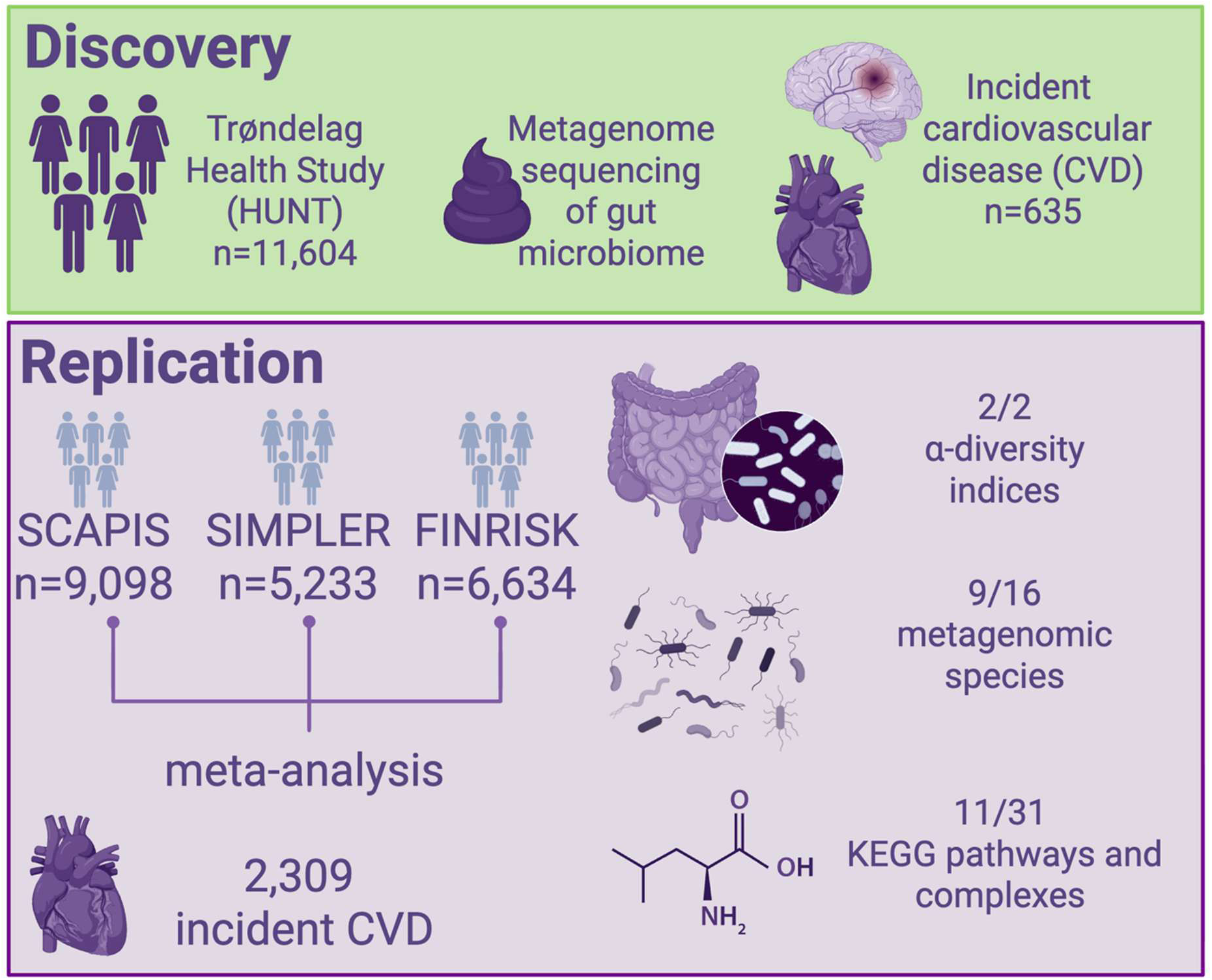
Discovery-replication framework used in four Nordic cohorts. After identifying gut microbiome features associated with incident CVD in the HUNT study as our discovery cohort, we replicated both α-diversity indices, 9 of 16 metagenomic species, and 11 of 31 Kyoto encyclopedia of genes and genomes (KEGG) modules in an inverse variance weighted fixed-effect meta-analysis of SCAPIS, SIMPLER, and FINRISK.

**Table 1.** Cohort descriptions. Baseline and summary statistics for the discovery cohort, HUNT study, and the replication cohorts—SCAPIS, SIMPLER, and FINRISK. HUNT, The Trøndelag Health Study; SD, standard deviation; SIMPLER, Swedish Infrastructure for Medical Population-Based Life-Course and Environmental Research; SCAPIS, Swedish CArdioPulmonary bioImage Study * = for FINRISK antibiotic prescription in the past 6 months ** = for FINRISK PPI prescription in the past 4 months

| Characteristic | Discovery cohort | Replication cohorts |  |  |
|---|---|---|---|---|
|  | HUNT | SCAPIS | SIMPLER | FINRISK |
| Number of participants | 11,604 | 9,098 | 5,233 | 6,634 |
| Baseline age, median (SD) | 60.90 (14.20) | 57.50 (4.35) | 73.17 (5.25) | 49.77 (12.74) |
| Number of females (%) | 7337 (63.23%) | 4,839 (53.19%) | 2715 (51.90%) | 3685 (55.55%) |
| Years of follow-up, median (SD) | 5.21 (0.80) | 7.64 (1.20) | 7.62 (2.62) | 17.82 (4.09) |
| Number of CVD (%) | 635 (5.47%) | 217 (2.38%) | 935 (17.87%) | 1157 (17.44%) |
| Number of deaths from other causes (censored) (%) | 305 (2.63%) | 165 (1.81%) | 683 (13.05%) | 417 (6.29%) |
| Number of current smokers | 905 (7.80%) | 1,231 (13.53%) | 260 (5.12%) | 1577 (23.77%) |
| Number of participants with antibiotic prescription* in past 3 months (%) | 919 (7.92%) | 567 (6.23%) | 328 (6.27%) | 1144 (17.24%) |
| Number of participants with proton pump inhibitor prescription** in past 3 months (%) | 1089 (9.38%) | 580 (6.38%) | 528 (10.09%) | 162 (2.44%) |
| Years of stool sample collection | 2018-2019 | 2014-2018 | 2011-2021 | 2002 |
| Date follow-up ended for diagnosis codes and mortality | 2023-12-31 | 2024-09-30 | 2024-12-31 | 2020-12-31 |
| Location | Trøndelag county, Norway | Uppsala and Malmö, Sweden | 3 counties in central Sweden | 6 regions in Finland |
| Average metagenomic sequencing read depth per sample | 22.9M | 25.3M and 26.3M | 51M | 900,000 |

### Outcome definition

In HUNT, incident cardiovascular disease (CVD) was defined as the first hospital diagnosis of ischemic heart disease (I20-I25), ischemic stroke (I63), or heart failure (I50), or cardiovascular death (I20-25, I63, I50), occurring after baseline (Supplementary Table 1). Participants with prevalent CVD at baseline, identified using the same ICD codes as those defining the outcome, were excluded. Participants were followed until the incident CVD diagnosis or CVD-related death, non-CVD related death, emigration, or end of follow-up on December 31, 2023. During follow-up, 635 incident CVD events were identified among 11,604 participants free from CVD at baseline. Cardiovascular deaths were identified from the Norwegian Cause of Death Registry (Dødsårsaksregisteret) using ICD codes for CVD listed as the underlying cause of death. Definitions of incident CVD were harmonized across replication cohorts to the extent possible (Supplementary Table 1). Hospital discharge diagnoses in the HUNT catchment area have been validated and show excellent validity (>80%) for stroke and acute myocardial infarction and moderate positive predictive value for heart failure (67%)^31^

### Metagenomics

In the HUNT study, stool sample handling, DNA extraction, library construction, and metagenomic sequencing were carried out according to established protocols described previously^32^. Briefly, 56,042 individuals participated in HUNT4 (2017-2019) and 13,268 provided stool samples. 50% of these have participated in all HUNT surveys. After sequencing and quality control, 12,887 individuals were included in this analysis. Samples were processed, and deep metagenomic sequencing was performed by Cmbio (Copenhagen, Denmark; previously named Clinical Microbiomics A/S). Three circular punches (6 mm diameter) were taken from each stool collection card and placed into designated wells of MagMAX™ 96 Deep Well Plates (Thermo Fisher Scientific, Waltham, MA, USA). Microbial cells were lysed by bead beating, after which DNA was purified using the MagMAX Ultra Microbiome kit (Thermo Fisher Scientific) in accordance with the manufacturer’s instructions on a KingFisher™ Flex instrument (Thermo Fisher Scientific). DNA was enzymatically fragmented and sequencing libraries were generated using the Celero EZ DNA-seq Core Module on a Tecan DreamPrep NGS automated platform.

Sequencing was performed on an Illumina NovaSeq 6000 system with paired-end 150 bp reads, producing an average depth of 22.9 million read pairs per sample with more than 85% of bases achieving a Phred quality score above 30. Metagenomics methodology in SCAPIS, SIMPLER, and FINRISK is described in the Supplementary Methods. Briefly, for SCAPIS, metagenomic sequencing using the Illumina Novaseq 6000 system yielded an average sequence depth of 25.3 million read pairs for the Uppsala samples and 26.3 million read pairs for Malmö samples. For SIMPLER, sequencing on MGI DNBSEQ G400 or T7 platforms generated an average of 51 million reads per sample. For FINRISK, sequencing on the Illumina HiSeq 4000 platform yielded an average of 900,000 reads per sample.

### Definition of diversity indices

To estimate α-diversity, abundance tables were first rarefied by randomly subsampling an equal number of signature gene counts from each sample without replacement. Species profiles derived from these rarefied data were then used to calculate α-diversity indices, namely Shannon diversity index and species richness^33^, based on relative species abundances.

Analyses were completed using the diversity function from the *vegan* R package^34^ in all cohorts.

### Definition of metagenomic species

In HUNT, SCAPIS, and SIMPLER, taxonomic profiling was performed by Cmbio using the CHAMP profiler with the Human Microbiome Reference (HMR05) catalogue^35^ and annotated using GTDB-Tk release R214. In FINRISK, taxonomic profiling was conducted using the GTDB release R214. In the HUNT study, we tested for the association between the relative abundance of 546 species and incident CVD. We required that these metagenomic species had a prevalence of over 30% and the relative abundance was transformed using inverse rank normal transformation. In addition, we used the CLR transformation in sensitivity analyses.

### Definition of functional modules

In HUNT, SCAPIS, and SIMPLER, genes were mapped to Kyoto Encyclopedia of Gene and Genomes (KEGG) pathways^36^ using KEGG v78.2 by Cmbio. In the HUNT study, we tested for the association between the abundance of 461 KEGG modules with a prevalence over 30% and incident CVD. Associations for KEGG modules were not assessed in FINRISK due to shallow metagenomic sequencing.

### Statistical analysis in discovery and replication cohorts

We tested for the association between time to event of CVD and 1) two indices of microbiota α-diversity—Shannon diversity index and species richness; 2) relative abundance of metagenomic species; and 3) abundance of KEGG modules. We performed a Cox proportional hazards model using time to event with right censoring. The age at sampling, sex, and a binary variable recording use of antibiotics in the 3 months before sampling were used as covariates.

Individuals with CVD before baseline were excluded and only individuals without missing covariates were included (e.g., complete cases).

We sought external replication for significant species and KEGG modules with a False Discovery Rate (FDR) adjusted p-value < 0.05 using the Benjamini-Hochberg procedure in the HUNT study, as well as both metrics of α-diversity. We performed a fixed-effect meta-analysis to combine effect sizes across SCAPIS, SIMPLER and FINRISK for α-diversity and species associations and across SCAPIS and SIMPLER for KEGG modules. We tested for heterogeneity between cohorts with Cochran’s Q-test and α of 5%.

We considered a finding to be replicated for each α-diversity index if the two-sided meta-analysis p-value was significant at an α of 5%. For species and KEGG modules, we considered a finding to be replicated if the meta-analysis estimate had the same direction of effect as the discovery analysis and reached significance in a one-sided test at a Bonferroni-corrected threshold of α =0.05/k where k is the number of features tested. Because the direction of effect was prespecified, this corresponds to a two-sided p-value <0.10/k, specifically p<0.0062 for species and p<0.00323 for KEGG. Proportional hazards assumptions were checked using the cox.zph function in R. After considering corrections for multiple testing, all model assumptions were met (Supplementary Table 2).

Analyses for the HUNT, SCAPIS, and FINRISK cohorts, as well as all meta-analyses and data visualization, were performed in R (versions 4.3.1-4.4.2). Analyses for the SIMPLER cohort were conducted in Stata (version 15.1; StataCorp, College Station, TX, USA). The R package pheatmap^37^ was used for creating heatmaps. Pearson correlations between the effect sizes for the KEGG module and MGS were estimated, and hierarchical clustering with complete linkage and Euclidean distance was used to group the modules and species based on the correlations.

### Sex differences

To explore whether the associations between 1*) Rothia mucilaginosa_A* and 2) manganese transport system KEGG module and CVD differed by sex, we added a multiplicative interaction term between sex and the exposure of interest to the primary models, with all other covariates unchanged. Interaction was assessed with a Wald test of the interaction coefficient. We then repeated the primary analyses stratified by sex, with sex removed from the covariate set.

### Sensitivity analysis

In the HUNT study, we performed several sensitivity analyses for the associations passing the FDR threshold in the discovery analysis and both metrics of α-diversity. We tested the impact of excluding all samples with antibiotic use, adjusting for the Bristol stool chart, excluding follow-up time < 1 year, and using CLR transformation. We also adjusted for self-reported diabetes, current use of antihypertensive medication and cholesterol lowering medication, use of diabetes medication including tablets, non-insulin injectables, or insulin, and combinations of these. In all four cohorts we performed sensitivity analyses to also adjust for current smoking, alcohol consumption, and proton pump inhibitor treatment within 3 months of sampling (Model 2), and for Model 2 covariates plus dietary variables (Model 3). Dietary variables and categories of alcohol consumption were harmonized across cohorts to the extent possible as described in the Supplementary Methods.

### Correlations with biomarkers in the HUNT study

To explore the correlations between the relative abundance of species and abundance of KEGG modules with cardiometabolic markers, we analyzed data on body mass index (BMI), waist-hip-ratio (WHR), systolic blood pressure (SBP), hemoglobin A1c (HbA1c), and non-fasting serum levels of triglycerides (TG), non-high density lipoprotein (non-HDL) cholesterol, and C-reactive protein (CRP). Partial Spearman correlations (R package *ppcor*^38^, v1.1) were estimated for the relative abundance of species or abundance of KEGG modules, adjusting for age, sex, and recent antibiotic use. Sex-stratified analyses adjusting only for age and antibiotic were also performed. For SBP, values were adjusted by adding 15 mmHg for individuals who self-reported antihypertensive medication use. For non-HDL cholesterol, the difference between total cholesterol and high-density lipoprotein cholesterol, the total cholesterol levels were divided by 0.8 for individuals who reported lipid-lowering medication use. For HbA1c, individuals who reported a history of diabetes were excluded.

### Two-sample Mendelian Randomization

We used denture-wearing as a proxy to assess the causal effect of severe tooth loss and advanced dental disease on the relative abundance of *Rothia mucilaginosa_A* in the gut. We employed a two-sample Mendelian randomization (MR) framework. SNPs serving as instrumental variables (IVs) for the exposure were drawn from the largest available GWAS of self-reported denture use, which combined data from the UK Biobank (UKB) and the Gene-Lifestyle Interactions in Dental Endpoints (GLIDE) consortium^39^. Summary statistics for *Rothia mucilaginosa_A* came from human genetic associations with relative abundance of gut microbiota species in the HUNT study^32^. Instrumental variants are listed in Supplementary Table 3.

As the primary MR analysis, we used inverse-variance weighted (IVW) estimates with fixed effect, as Cochran’s Q test indicated no significant heterogeneity among the genetic instruments. MR-Egger regression was performed as a sensitivity analysis to assess potential directional horizontal pleiotropy^40^. To further account for possible heterogeneity and outlier effects among SNPs, we applied MR-PRESSO to identify and correct for outlying genetic instruments^41^. In addition, weighted median MR was conducted as a complementary sensitivity analysis. All MR analyses were performed in R using the *MendelianRandomization*^42^ and *MRPRESSO*^41^ packages.

### Correlations with metabolites in SCAPIS and denture use in SIMPLER

For the 9 replicated species, we performed partial Spearman correlations for the plasma metabolome similar to what is previously described, but with the updated CHAMP profiler for the species^43^. In brief, metabolites detected in <100 participants were removed, metabolites detected in <2% of participants were considered as present or absent, and drug metabolites were considered as present or absent with the exception of salicylate and 2,6-dihydroxybenzoic acid, which were present in >99% of individuals. For each metabolite, values below the detection threshold were imputed by the minimum value above the detection threshold, and the results were batch-normalized. Place of birth was categorized as Sweden, Non-Swedish Europe, and other. Covariates were age, sex, place of birth, microbial DNA extraction plate, metabolomics delivery batch, and antibiotic use (defined in SCAPIS as dispensed prescription in the past 6 months). We present the top 5 metabolites per replicated species. Denture use in SIMPLER was self-reported from the question “Do you use a removable partial or full dental prosthesis (denture) to replace missing teeth?” Linear regression for relative abundance of *Rothia mucilaginosa_A* with denture use was adjusted for age, sex, antibiotic use (defined in SIMPLER as dispensed prescription in the last 3 months), proton pump inhibitor use in the last 3 months, body mass index, diabetes, and smoking.

### Ethics statement

All procedures were conducted in accordance with the Declaration of Helsinki on ethical principles for medical research involving human participants. All participants in HUNT2-4 provided written informed consent. The HUNT cohort was approved by the Regional Committee for Medical and Health Research Ethics, Central Norway (REK-656785). The analyses of SIMPLER data were approved by the Swedish Ethical Review Authority (Dnr 2019-03986 with amendment Dnr 2024-04685-02). The FINRISK 2002 study was approved by the Coordinating Ethical Committee of the Helsinki and Uusimaa University Hospital District (REF 558/E3/2001). All participants provided written informed consent. The Swedish Ethical Review Authority has approved the studies in SCAPIS (DNR 2022-06137-01 with amendment 2025-03082-02) and all included participants have provided informed consent. The data used in this manuscript is not publicly available due to legal and privacy limitations.

## Results

We applied a discovery-replication framework to identify reproducible gut microbiome signatures associated with CVD (Figure 1). Among 11,604 participants in the HUNT discovery cohort, we identified 635 incident CVD cases (5%) during a median follow-up of 5.2 years. The incidence of CVD differed across the three replication cohorts, with the highest incidence observed in SIMPLER (935 cases; 18%) and FINRISK (1,157 cases; 17%), reflecting the higher baseline age in SIMPLER and the longer follow-up time in FINRISK, whereas the lowest incidence was observed in SCAPIS (217 cases; 2%), attributable to its lower baseline age, shorter follow-up time, and the omission of heart failure from the CVD definition (Table 1).

### Associations with α-diversity

Higher gut microbiota α-diversity was consistently associated with a lower risk of CVD (Table 2, Supplementary Table 4). In the HUNT study, each standard deviation (SD) increase in the Shannon diversity index was associated with a 9% lower hazard of CVD (HR=0.91; 95% CI 0.84-0.99). Similarly, higher species richness was associated with a lower risk of CVD (HR=0.91; 95% CI 0.84-0.99). In a meta-analysis of the three replication cohorts (SCAPIS, SIMPLER, and FINRISK; Table 2, Supplementary Table 4, Supplementary Figure 1), increases in both the Shannon diversity index (HR=0.90; 95% CI 0.86–0.94) and species richness (HR=0.88; 95% CI 0.85–0.92) were associated with a reduced hazard of incident CVD, with effect estimates closely mirroring those observed in HUNT. There was no significant heterogeneity across the three replication cohorts (Supplementary Table 4).

**Table 2.** Replication for α-diversity indices. Greater α-diversity is associated with a decreased hazard of incident CVD in both the HUNT study (discovery) and the fixed-effect meta-analysis of SCAPIS, SIMPLER, and FINRISK (replication). HR=hazard ratio, CI=confidence interval. HRs are given per standard deviation increase in α-diversity metrics.

|  | **Discovery cohort**<br>**(The HUNT Study)** |  | **Replication meta-analysis**<br>**(SCAPIS, SIMPLER, FINRISK)** |  |
|---|---|---|---|---|
| **Index** | **HR (95% CI)** | **p-value** | **HR (95% CI)** | **p-value** |
| **Shannon Index** | 0.91 (0.84-0.99) | 2.3x10<sup>-2</sup> | 0.90 (0.86-0.94) | 1.1x10<sup>-6</sup> |
| **Richness** | 0.91 (0.84-0.99) | 3.2x10<sup>-2</sup> | 0.88 (0.85-0.92) | 1.8x10<sup>-8</sup> |

### Associations with metagenomic species

In the HUNT study we identified 16 species associated with incident CVD (5% FDR, Supplementary Table 5, Supplementary Figure 2A). In a meta-analysis of the three replication cohorts, nine of the sixteen species met the predefined statistical significance threshold for replication (Figure 2A, Supplementary Figure 3, Supplementary Table 6), with all nine species exhibiting a consistent direction of effect compared with the discovery cohort. For the replicating species with the lowest p-value, *Blautia_A caecimuris,* a 1-SD increase in relative abundance was associated with 15% higher hazard of incident CVD (HR=1.15; 95% CI 1.09-1.20). *Rothia mucilaginosa_A, Mediterraneibacter torques,* and *Thomasclavelia ramosa* were also significantly associated with a higher hazard of incident CVD, whereas *Egerieenecus* sp900553315, UBA11524 sp000437595*, Blautia_A* sp900066205, UMGS882 sp900546385, and *Marvinbryantia* sp900066075 were associated with a decreased hazard of incident CVD in both the discovery and replication cohorts (Figure 2A, Supplementary Figure 3, Supplementary Table 6). There was no significant heterogeneity across the three replication cohorts (Supplementary Table 6).

**Figure 2.**
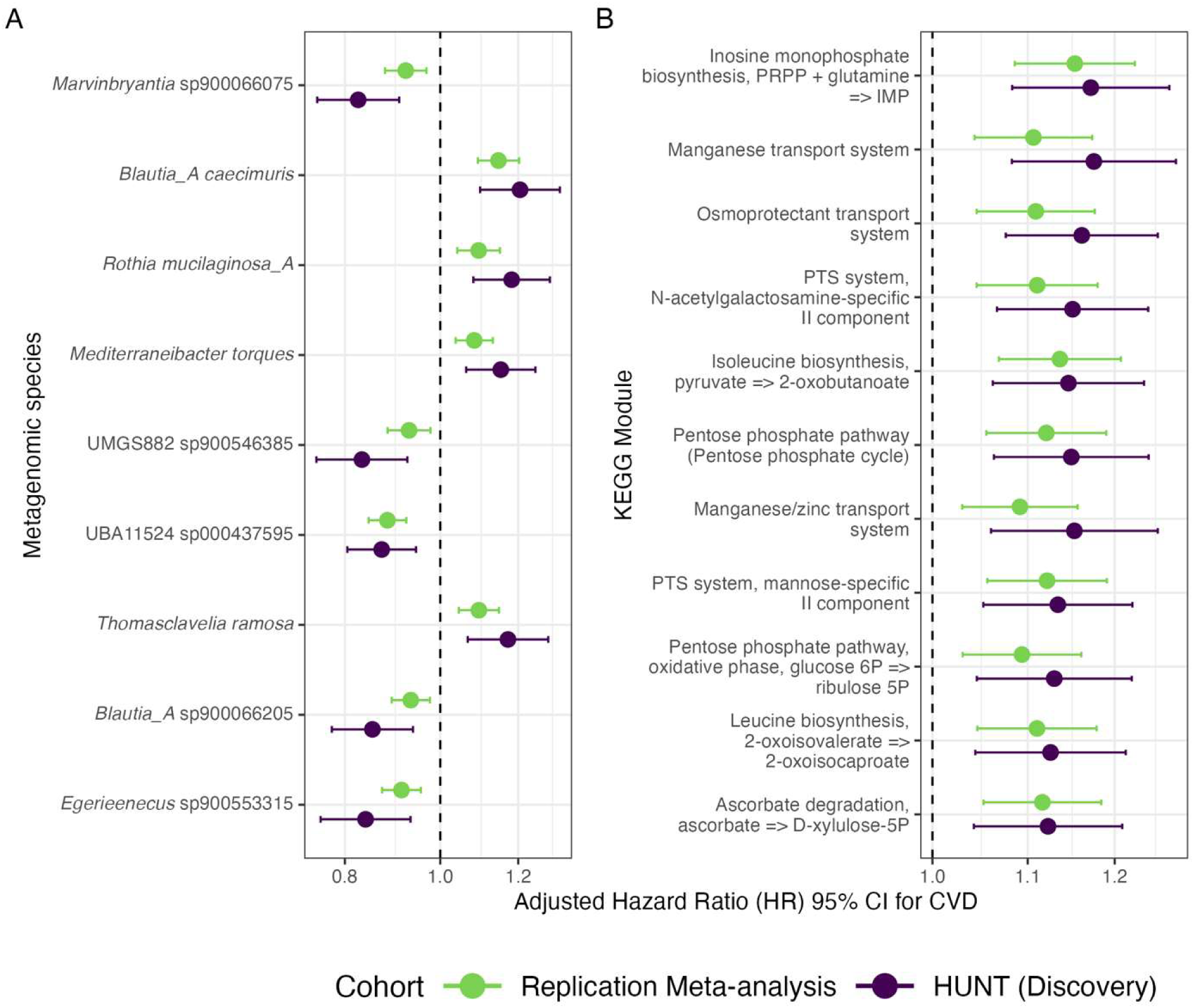
Gut microbial species and functional KEGG modules associated with incident CVD in discovery and replication cohorts. (A) Estimated effect sizes for the 9 replicated metagenomic species in the HUNT study (discovery) and a fixed-effect meta-analysis with inverse variance weighting of the replication cohorts SCAPIS, SIMPLER, and FINRISK. Species are ordered from most to least significant by the discovery analysis p-value. Hazard ratios are given per standard deviation increase in relative abundance of species. (B) Estimated effect sizes for the 11 replicated KEGG modules in the HUNT study (discovery) and a fixed-effect meta-analysis of SCAPIS and SIMPLER (replication). Modules are ordered from most to least significant by the discovery analysis p-value. Hazard ratios are given per standard deviation increase in abundance of KEGG modules.

### Pathways underlying the association between gut microbiota and incident CVD

In the HUNT study, we identified 31 functional KEGG modules associated with incident CVD that remained significant at a 5% FDR (Supplementary Figure 2B, Supplementary Table 7). In a meta-analysis of the two replication cohorts with KEGG data available (SIMPLER, SCAPIS), we replicated 11 of these 31 KEGG modules (Figure 2B, Supplementary Figure 4, Supplementary Table 8). All replicated KEGG pathways were positively associated with incident CVD. A 1-SD increase in the abundance of the most statistically significant KEGG pathway, *“Inosine monophosphate biosynthesis, PRPP + glutamine => IMP”*, was associated with a 17% increased hazard of incident CVD in the HUNT study (HR=1.17; 95% CI 1.08-1.27) and a 15% higher hazard in the replication meta-analysis (HR=1.15; 95% CI 1.09-1.22). For the eleven replicated KEGG modules, the effect estimates were in general comparable between the HUNT discovery cohort and the meta-analyzed replication cohorts, and no significant heterogeneity was observed between the SCAPIS and SIMPLER replication cohorts (Figure 2B, Supplementary Figure 4, Supplementary Table 8).

Correlation analysis between replicated species and replicated functional KEGG modules revealed two general patterns. The first pattern was that the relative abundance of *Rothia mucilaginosa_A* was positively associated with the abundances of two KEGG modules involved in manganese transport systems, and higher levels of these modules were in turn associated with an increased hazard of incident CVD (Figure 3A, Supplementary Table 9). The second, and more common, pattern was that the relative abundances of all other replicated species associated with increased risk of CVD were positively associated with the remaining replicated KEGG modules, whereas species associated with reduced risk of incident CVD were negatively associated with these modules (Figure 3A, Supplementary Table 9).

**Figure 3.**
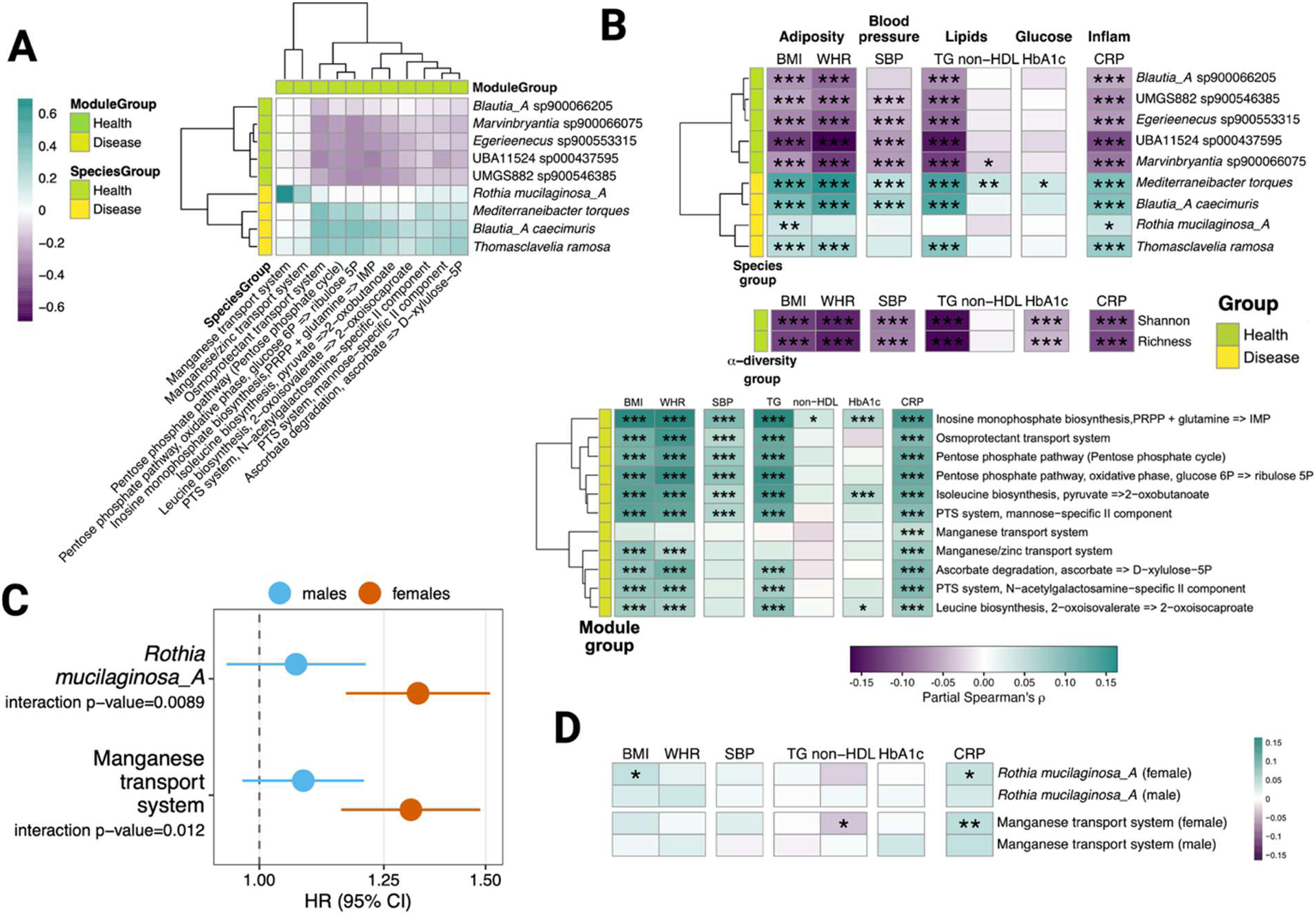
Correlations between microbial features and biomarkers in the HUNT study. (A) Heat map of the correlations between the nine replicated species and the eleven replicated functional KEGG modules. Spearman correlations (rho) between the relative abundances of the species and the abundances of the KEGG modules in HUNT, presented as a heat map. Disease group for species (yellow) indicates that these species associate with higher risk of incident CVD while the health group (green) indicates species associated with lower risk of incident CVD. (B) Partial Spearman correlation (adjusted for age, sex, and antibiotic use) between α-diversity metrics, replicated species, and replicated KEGG modules and parameters reflecting the metabolic syndrome. * Bonferroni-adjusted p< 0.05, ** Bonferroni-adjusted p< 0.005, and *** Bonferroni-adjusted p< 0.0005. BMI = body mass index, WHR = waist-to-hip ratio, SBP = systolic blood pressure, TG = triglycerides, CRP = C-reactive protein. (C) Sex-stratified analyses of the associations between the relative abundance of *Rothia mucilaginosa_A* and the abundance of the KEGG module for the manganese transport system with incident CVD in the HUNT discovery cohort. Hazard ratios are presented per standard-deviation increase in the gut microbiome feature. D) Sex-stratified partial Spearman correlations (adjusted for age and antibiotic use) between the relative abundance of *Rothia mucilaginosa_A* and the abundance of the KEGG module for the manganese transport system, and parameters reflecting the metabolic syndrome. * Bonferroni-adjusted p< 0.05, ** Bonferroni-adjusted p< 0.005, and *** Bonferroni-adjusted p< 0.0005.

We next examined whether the gut microbiome features associated with incident CVD were also related to markers of the metabolic syndrome, a major determinant of cardiovascular risk, as assessed in the HUNT discovery cohort (Supplementary Table 10). Lower gut microbiota α-diversity (Shannon diversity index and species richness), associated with an increased risk of incident CVD, was strongly related to multiple markers of the metabolic syndrome, including elevated waist-to-hip ratio, increased blood pressure, high triglyceride levels, elevated HbA1c, and systemic inflammation as reflected by higher CRP concentrations (Figure 3B, Supplementary Table 11). Eight of the nine replicated species associated with incident CVD were also strongly linked to multiple markers of the metabolic syndrome in the expected direction, consistent with their relationship to incident CVD, with particularly pronounced associations with waist-to-hip ratio, triglyceride levels, blood pressure, and CRP (Figure 3B, Supplementary Table 11). These species were also often strongly associated with circulating metabolites reflecting secondary bile acid metabolism, androgenic steroid metabolism, and benzoate metabolism in the SCAPIS replication cohort (Supplementary Table 12), which could potentially link these gut microbiota–derived pathways to metabolic syndrome and incident CVD risk.

#### Rothia mucilaginosa

*A* was not associated with waist-to-hip ratio, triglyceride levels, or blood pressure, but showed an association with markers of systemic inflammation (Figure 3B, Supplementary Table 11). This pattern suggests that a distinct biological pathway, rather than traditional metabolic syndrome–related pathways, may underlie the link between this species and increased risk of CVD. This notion is further supported by the observation that the abundance of the manganese transport system, strongly and directly associated with the relative abundance of *Rothia mucilaginosa_A* (Figure 3A) and reproducibly linked to increased risk of incident CVD (Figure 2B), was likewise not associated with waist-to-hip ratio, triglyceride levels, or blood pressure, but was associated with systemic inflammation (Figure 3B, Supplementary Table 11).

To further dissect the underlying mechanism, we investigated the association between *Rothia mucilaginosa_A* and circulating metabolites in the SCAPIS replication cohort. Among the 964 circulating metabolites examined, ImP, a gut microbiota–derived metabolite closely linked to cardiovascular disease, was the metabolite most strongly associated with *Rothia mucilaginosa_A* (partial Spearman ρ = 0.15; *P* = 1.1 × 10^−38^; n = 8351; Supplementary Table 12).

The relative abundance of *Rothia mucilaginosa* is high in the oral cavity and correlates strongly with its levels in the gut^6^. Furthermore, denture use was associated with a higher relative abundance of *Rothia mucilaginosa_A* in the gut in the SIMPLER cohort (*β*^^^ = 0.19; 95% CI 0.12– 0.26; adjusted for age, sex, antibiotic use; n = 5,473). The association is almost identical after further adjustment for recent PPI use (*β*^^^ = 0.25; 95% CI 0.16-0.33) and is only slightly attenuated after additional adjustment for BMI, diabetes, and smoking (*β*^^^ = 0.21; 95% CI 0.13-0.29). Finally, Mendelian randomization revealed that genetically predicted denture use was causally associated with higher relative abundance of *Rothia mucilaginosa_A* in the gut (β= 0.25; 95% CI 0.03-0.47, Supplementary Figure 5, Supplementary Table 13). Similar estimates were observed with the weighted median Mendelian randomization approach, and there was no evidence of directional pleiotropy (MR-Egger intercept p=0.43), no heterogeneity (Cochran’s Q p=0.23), and no outlier variants detected by MR-PRESSO (Supplementary Table 13).

Thus, for *Rothia mucilaginosa A*, a prevalent oral-derived gut microbiota species linked to poor oral health, a pathway linking this taxon to increased risk of incident CVD emerges, linked to altered gut microbial manganese transport, elevated circulating levels of ImP, and systemic inflammation (Figure 4). Previous studies have shown that ImP promotes atherosclerosis and is associated with endothelial dysfunction, mechanisms that may contribute to increased risk of CVD^10,11^ (Figure 4).

**Figure 4.**
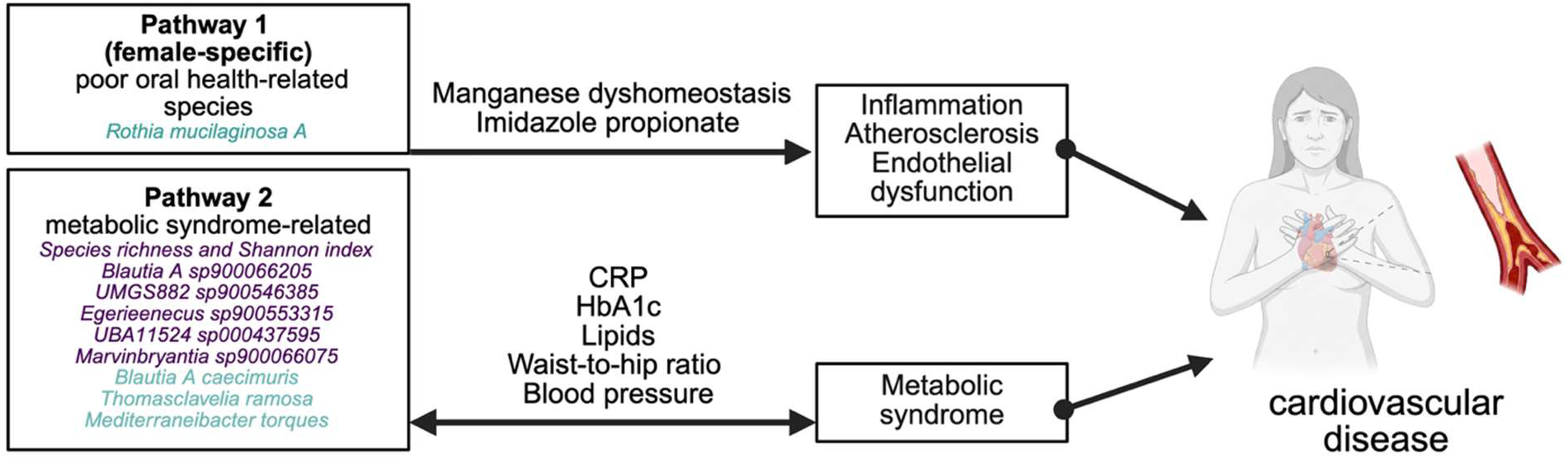
Proposed pathways underlying the association between gut microbiota and incident CVD. Two main proposed pathways for the link between gut microbiota and incident CVD. Associations linked to increased risk of CVD are in blue and decreased risk of CVD are in purple. A first female-specific pathway involves *Rothia mucilaginosa_A*, a prevalent oral gut microbiota species, which was enriched among denture users, serving as a proxy for longstanding poor oral health. This species is linked to increased risk of incident CVD through affected gut microbial manganese transport, elevated circulating concentrations of imidazole propionate and systemic inflammation. The second pathway involves α-diversity metrics and eight replicated species, all of which were strongly associated with multiple markers of the metabolic syndrome, an established major determinant of incident CVD.

As a previous report showed that high *Rothia mucilaginosa* associates with higher coronary artery calcification score with a significant sex interaction^6^, we performed sex-stratified analyses of the association between *Rothia mucilaginosa_A* and incident CVD (Supplementary Table 14). *Rothia mucilaginosa_A* was associated with incident CVD in females (HR=1.33; 95% CI 1.17-1.51) but not males (HR=1.07; 95% CI 0.94-1.21) with a significant interaction with sex (p= 0.0089; Figure 3C, Supplementary Table 14). Similarly, for the functional KEGG module manganese transport system, which was strongly correlated with *Rothia mucilaginosa A*, we observed a significant association in females (HR=1.31; 95% CI 1.16–1.49) but not in males (HR=1.08; 95% CI 0.97–1.21) with a significant interaction with sex (p=0.012; Figure 3C, Supplementary Table 14). Furthermore, the correlations of both *Rothia mucilaginosa_A* and the manganese transporter system with CRP appeared slightly more pronounced in females than in males (Figure 3D, Supplementary Table 15). We found no association between circulating estradiol or testosterone and the relative abundance of *Rothia mucilaginosa_A* in females, indicating that sex steroids do not influence its abundance (Supplementary Table 16).

### Sensitivity analyses

In the HUNT discovery study, associations for α-diversity metrics (Supplementary Table 17), species (Supplementary Table 18), and KEGG modules (Supplementary Table 19) with incident CVD remained largely unchanged after adjustment for Bristol stool scale (a proxy for bowel motility), exclusion of participants on antibiotic treatment within 3 months of sampling, removal of participants with an event in the first year of follow-up and when using CLR transformation. In the HUNT study, both diversity indices were no longer significant after adjusting for combinations of diabetes and medications for diabetes, cholesterol, and blood pressure, but the hazard ratios remained similar between 0.92 and 0.94 (Supplementary Table 17). However, all metagenomic species and KEGG module associations that passed the FDR threshold in the discovery analysis remained nominally significant (p-value <0.05) with very similar hazard ratio estimates after adjusting for combinations of diabetes and medications (Supplementary Table 18-19). In general, the effect estimates were robust to further adjustment for alcohol consumption, current smoking status, and use of proton pump inhibitors (Model 2) and diet (Model 3) both in the discovery cohort and in the replication meta-analysis (Supplementary Figures 1, 3, 4, Supplementary Tables 20-25).

## Discussion

Prior studies have linked gut microbiota to cardiovascular health, but most were cross-sectional, small, used 16S rRNA sequencing, and/or unreplicated^3,17,44–48^. Thus, microbial features consistently associated with incident CVD remain insufficiently defined. In the present study, we examined stool-derived metagenomes within a discovery–replication framework across four large Nordic cohorts (N=32,569; n=2,944 incident CVD events across the four cohorts). We identified the first gut microbiome features reproducibly associated with incident CVD, including two indices of α-diversity, nine species, and eleven functional KEGG modules. We observed two main pathways for these associations between the gut microbiome and incident CVD. The first pathway involved the prevalent oral–gut species *Rothia mucilaginosa_A*, linking it to increased incident CVD risk through affected gut microbial manganese transport, elevated circulating ImP, and systemic inflammation, and this relationship appeared to be female-specific. The second pathway involved α-diversity metrics and the remaining eight species, which were strongly linked to metabolic syndrome. Collectively, these results highlight two principal pathways through which the gut microbiome may be linked to CVD (Figure 4).

CVD incidence and severity differ markedly by sex and gender^49,50^. There is a clear protection against CVD in premenopausal women, linked to the beneficial effects of estrogens in preventing atherosclerosis^51^. As a result, men display CVD at younger ages than women, with an estimation of approximately 7-years earlier development^52^. However, women experience a substantial post-menopausal rise in CVD. Beyond sex steroids, there remains an important gap in female-specific biomarkers and pathways for cardiovascular disease^53,54^. Notably, we found that *Rothia mucilaginosa_A* was associated with incident CVD exclusively in females. Moreover, *Rothia mucilaginosa_A* showed no associations with waist-to-hip ratio, triglyceride levels, or blood pressure, while demonstrating a link with systemic inflammation, suggesting that a distinct biological pathway, rather than traditional metabolic syndrome-related pathways, may underlie its association with increased CVD risk in females.

Furthermore, the abundance of the functional KEGG module for manganese transport system, which was strongly correlated with *Rothia mucilaginosa_A*, was also linked to increased risk of incident CVD specifically in females, while showing no associations with metabolic-syndrome– related parameters. Manganese is a cofactor that protects against oxidative stress^55^ and may be involved in competitive dynamics in the gut microbiome^56^. Manganese homeostasis is an essential component of the host-pathogen interface^57^ with dietary manganese protecting bacteria from the immune system in a mouse model of bacterial endocarditis^58^. Circulating manganese has been reported to be associated with cardiovascular disease, with some evidence suggesting that these associations may be more pronounced in women^59–61^. Further studies are required to clarify the relevance of gut microbiota–mediated manganese transport for CVD in women.

Given the observational and causal evidence that denture use, serving as a proxy for longstanding poor oral health, is associated with the relative abundance of *Rothia mucilaginosa*_A, these findings raise the possibility that this species may, at least in part, mediate the previously reported association between poor oral health and CVD^62^. Further studies are needed to clarify this potential pathway and its implications for CVD prevention. Interestingly, circulating ImP, a gut microbiota–derived metabolite closely linked to CVD^10,11^, was the metabolite most strongly associated with relative abundance of *Rothia mucilaginosa_A*. ImP has been proposed to drive atherosclerosis through the imidazoline-1 receptor (I1R) in myeloid cells, and pharmacological blockade of ImP binding to I1R inhibited atherosclerotic progression and inflammation in mice, identifying the ImP-I1R axis as a potential therapeutic target^63^. Thus, for *Rothia mucilaginosa_A*, a prevalent oral gut microbiota species, a female-specific pathway linking this species to increased risk of incident CVD emerges that may involve affected manganese handling, elevated circulating ImP, and systemic inflammation (Figure 4). Although we consider a gut-derived pathway the most plausible explanation, we cannot exclude that *Rothia mucilaginosa_A* may also exert direct adverse cardiovascular effects. This notion is supported by reports describing the species as an opportunistic pathogen capable of hematogenous spread in vulnerable populations^64^ and documented presence in endocarditic lesions^65,66^.

The second pattern linked incident CVD to gut microbiome parameters, including α-diversity metrics and eight species which were also strongly linked to multiple metabolic-syndrome markers in the expected direction. Low α-diversity, reflected by reduced Shannon diversity index and species richness, was associated with a higher risk of CVD as well as elevated waist-to-hip ratio, systolic blood pressure, triglycerides, HbA1c, and CRP. These are characteristic features of the metabolic syndrome which is closely linked to increased cardiovascular risk^67,68^. This metabolic syndrome-associated pathway was also associated with several functional gut microbiota-associated pathways reflecting purine metabolism, microbial stress responses, altered redox balance, ascorbic acid degradation, and branched-chain amino acid biosynthesis.

Our observation that lower Shannon diversity index and species richness were consistently associated with a higher risk of incident CVD aligns with prior cross-sectional findings from the Framingham Heart Study, demonstrating that decreased gut microbiota α-diversity was associated with an increased calculated 10-year risk of CVD^3^. We also observed that low α-diversity was associated with high CRP, reflecting systemic inflammation. Collectively, these findings underscore that reduced gut microbiota α-diversity, a signature of the metabolic syndrome and an unstable and inflammation-prone gut ecosystem, marks increased cardiovascular risk.

*Marvinbryantia* sp900066075, the metabolic-syndrome–related species most strongly associated with reduced risk of incident CVD in the discovery cohort, belongs to a genus positively associated with dietary fiber intake, favorable metabolic trait profiles, and lower odds of type 2 diabetes^69^. This species remained significantly associated with CVD after adjusting for diet. The only formally characterized species in the genus is *Marvinbryantia formatexigens* which ferments carbohydrates to the short-chain fatty acid acetate. Two of the species identified here, *UBA11524* sp000437595 and *Blautia*_A sp900066205 were recently associated with glomerular filtration rate^70^, suggesting a potential link between gut microbial composition, renal function, and cardiovascular health.

The most significant metabolic syndrome-related and replicated functional KEGG module was the inosine monophosphate (IMP) biosynthesis pathway which creates a key precursor for building blocks of DNA, RNA, and cellular energy. Microbes with affected purine biosynthetic capacity may contribute to pro-inflammatory and pro-oxidative host environments relevant to CVD through metabolites generated in this pathway, including uric acid, its terminal product^71^. The association between elevated abundance of the microbial osmoprotectant systems and increased CVD risk suggests adaptation to hyperosmotic stress, which may arise during dehydration, high salt intake, or intestinal inflammation. This may contribute to cardiometabolic risk by promoting metabolic dysregulation and systemic inflammation and by reflecting a gut microbial stress phenotype linked to atherogenesis^72^. These associations for functional KEGG modules with incident CVD suggest gut microbial features that may promote low-grade chronic inflammation, providing a biological basis for atherosclerotic plaque progression and thrombosis and thereby contributing to CVD.

We observed that increased relative abundance of *Mediterraneibacter torques* was associated with increased risk of CVD and was significantly positively correlated with all tested metabolic parameters. This is consistent with previous studies linking *Mediterraneibacter torques* to high blood pressure and adiposity^73,74^. In addition, *Mediterraneibacter torques* is a specialist in mucin degradation and has been associated with intestinal inflammation, potentially driven by increased gut permeability resulting from reduced mucin-protective layers, thereby promoting immune activation and systemic inflammation.

Our study has several strengths. It is by far the largest investigation conducted to date and includes a successful replication phase with complete follow-up enabled through linkage to high-quality Nordic registers. We used metagenomic sequencing rather than 16S rRNA gene sequencing, which enables species-level resolution and functional characterization. Because we were able to perform sensitivity analyses accounting for diet as well as proton pump inhibitor and antibiotic use, we could adjust for notable confounders. Finally, our study was conducted in populations with deep phenotyping, enabling the investigation of potential underlying mechanisms.

Our findings should be interpreted in the context of the study limitations. First, our analysis only indicates microbiome features correlated with, but not necessarily causal for, CVD. We rely on diagnosis codes from hospitals and death registries, rather than validated outcomes, which may be imperfect and result in misclassification. Furthermore, only prescribed medications are captured in the prescription databases, so the binary PPI exposure variable may underestimate true PPI usage. Although metagenomic sequencing allows species-level resolution, some disease-causing metabolic activities of the gut microbiome may exist at the strain level, which we were unable to account for. Residual confounding by unmeasured factors or by imprecise measurement of adjusted covariates such as self-reported diet, remains possible. Finally, our findings may not be generalizable to other global populations outside of the Nordic region.

The use of the CVD outcome has advantages and disadvantages. The composite CVD outcome combines cardiovascular outcomes with partially distinct pathophysiology. While myocardial infarction and stroke are closely related to atherothrombosis and plaque instability, heart failure reflects more heterogeneous mechanisms, from arrhythmias to valvular disease. However, the composite CVD endpoint is frequently used in randomized clinical trials, so our findings are informative for future interventional studies. In addition, combining the cardiovascular outcomes into a composite outcome likely aided in statistical power to identify associations. While the CVD definition was harmonized across cohorts, there was some heterogeneity, for example, data for heart failure was not available for SCAPIS.

In conclusion, we identified gut microbiome features reproducibly associated with incident CVD. We observed two main pathways for these associations between gut microbiota and incident CVD. The first pathway was female-specific and involved the oral-gut species *Rothia mucilaginosa*_A, which was enriched among denture users, serving as a proxy for longstanding poor oral health. This pathway linked *Rothia mucilaginosa_A* to increased incident CVD risk through altered manganese transport, elevated circulating ImP levels, and heightened systemic inflammation. (Figure 4). The second more general pathway involved α-diversity metrics and eight species, which were strongly associated with multiple markers of the metabolic syndrome. These findings indicate that gut microbiota diversity, specific taxa, and functional pathways may provide new insights into the etiology of CVD and may ultimately help guide its prevention and treatment. Furthermore, these findings suggest that the link between poor oral health and CVD may, at least partly, operate through altered gut microbiota.

## Acknowledgements

B.N.W., K.H. and C.O. planned and designed the study. K.H. and C.O. collected data from HUNT; S.C.L. and K.M. collected data from SIMPLER; T.N., V.S., R.K., and A.S.H. collected data from FINRISK; J.Ä., M.O.-M., J.G.S, G.E. and T.F collected data from SCAPIS; C.G.V. and G.M., performed the taxonomic profiling for FINRISK; M.N., E.C, L.G. and L.L. analyzed HUNT data; S.C.L. analyzed SIMPLER data; K.P. and K.F.D analyzed SCAPIS data, O.K. analyzed FINRISK data; B.N.W. drafted the analysis plan and performed the meta-analyses; B.N.W. wrote the first draft of the manuscript. All authors provided intellectual input, reviewed, and approved the final version of the manuscript. B.N.W, K.H., and C.O. are the guarantors of the study.

ChatGPT 5.1, Claude Opus 4.7, and Microsoft Copilot were used for summarizing and synthesizing internet and literature sources and drafting portions of text which were reviewed and revised by authors.

The Trøndelag Health Study (HUNT) is a collaboration between HUNT Research Centre (Faculty of Medicine and Health Sciences (MH), Norwegian University of Science and Technology (NTNU)), Trøndelag County Council, Central Norway Regional Health Authority, and the Norwegian Institute of Public Health.

We acknowledge SIMPLER for the data access, and we would like to thank Anna-Karin Kolseth and Niclas Håkansson for assistance.

The research in The Swedish CArdioPulmonary bioImage Study (SCAPIS) Resource, was conducted under Petition Number 774. We would like to acknowledge the help of Biobank Sweden and the local biobank facilities for their services in handling of biological samples and biobanking.

The FINRISK data used for the research were obtained from THL Biobank (study number: THLBB2025_17). We thank all FINRISK 2002 study participants for their generous participation in Finnish health research. We thank the Health and Social Data Permit Authority Findata for providing the health register data (permit number: THL/3273/14.02.00/2025). We wish to acknowledge CSC – IT Center for Science, Finland, for computational resources.

## Data Availability Statement

Due to the sensitive nature of the data and compliance with the General Data Protection Regulation, individual-level data cannot be made publicly available.

Individual level data from HUNT can be accessed by, or in collaboration with, a Norwegian principal investigator. Researchers can apply for HUNT data access from HUNT Research Centre (https://www.ntnu.edu/hunt) if they have obtained project approval from the Regional Committee for Medical and Health Research Ethics (REC). Information on the application and conditions for data access is available at https://www.ntnu.edu/hunt/data.

Data are available from SIMPLER (https://www.simpler4health.se) for researchers who meet the data access criteria, including ethical approval from the Swedish Ethical Review Authority and an approved data access application.

The FINRISK 2002 data are available from the Finnish Institute for Health and Welfare Biobank upon application (https://thl.fi/en/web/thl-biobank/for-researchers/application-process).

Access to SCAPIS data requires ethical approval from the Swedish Ethical Review Board and approval from the SCAPIS Data access board (https://www.scapis.org/data-access).

